# Estimating serum cross-neutralizing responses to SARS-CoV-2 Omicron sub-lineages elicited by pre-Omicron or Omicron breakthrough infection with exposure interval compensation modeling

**DOI:** 10.1101/2023.02.08.23285673

**Authors:** Sho Miyamoto, Yudai Kuroda, Takayuki Kanno, Akira Ueno, Nozomi Shiwa-Sudo, Naoko Iwata-Yoshikawa, Yusuke Sakai, Noriyo Nagata, Takeshi Arashiro, Akira Ainai, Saya Moriyama, Noriko Kishida, Shinji Watanabe, Kiyoko Nojima, Yohei Seki, Takuo Mizukami, Hideki Hasegawa, Hideki Ebihara, Shuetsu Fukushi, Yoshimasa Takahashi, Ken Maeda, Tadaki Suzuki

## Abstract

Understanding the differences in serum cross-neutralizing responses against SARS-CoV-2 variants, including Omicron sub-lineages BA.5, BA.2.75, and BQ.1.1, elicited by exposure to distinct antigens is essential for developing COVID-19 booster vaccines with enhanced cross-protection against antigenically distinct variants. However, fairly comparing the impact of breakthrough infection on serum neutralizing responses to several variants with distinct epidemic timing is challenging because responses after breakthrough infection are affected by the exposure interval between vaccination and infection. We assessed serum cross-neutralizing responses to SARS-CoV-2 variants, including Omicron sub-lineages, in individuals with breakthrough infections before or during the Omicron BA.1 epidemic. To understand the differences in serum cross-neutralizing responses after pre-Omicron or Omicron breakthrough infection, we used Bayesian hierarchical modeling to correct the cross-neutralizing responses for the exposure interval between vaccination and breakthrough infection. The exposure interval required to generate saturated cross-neutralizing potency against each variant differed by variant, with variants more antigenically distant from the ancestral strain requiring a longer interval. Additionally, Omicron breakthrough infection was estimated to have higher impact than booster vaccination and pre-Omicron breakthrough infection on inducing serum neutralizing responses to the ancestral strain and Omicron sub-lineages. However, the breadth of cross-neutralizing responses to Omicron sub-lineages, including BQ.1.1, after Omicron or pre-Omicron breakthrough infection with the ideal exposure interval were estimated to be comparable. Our results highlight the importance of optimizing the interval between vaccine doses for maximizing the breadth of cross-neutralizing activity elicited by booster vaccines with or without Omicron antigen.

**Significance Statement:** SARS-CoV-2 infections after vaccination with COVID-19 mRNA vaccines with the ancestral spike antigen induce high serum neutralizing responses against Omicron sub-lineages, which are antigenically distant from the ancestral antigen. In individuals with breakthrough infections, the exposure interval from vaccination to infection is critical for the induction of serum cross-neutralizing activity. We used statistical modeling to estimate the serum neutralizing response to Omicron sub-lineages corrected for the influence of different exposure intervals between vaccination and breakthrough infection in individuals with pre-Omicron and Omicron breakthrough infections. This enabled us to assess fairly the effects of exposure to distinct antigens on inducing serum cross-neutralizing responses with the ideal exposure interval, and revealed the clinical significance of optimizing the dose interval in COVID-19 booster vaccination.

## Introduction

At the end of 2021, the severe acute respiratory syndrome coronavirus 2 (SARS-CoV-2) Omicron (B.1.1.529) variant emerged and rapidly spread worldwide. Since then, the Omicron variant has evolved into multiple sub-lineages, with BA.1, BA.2, and BA.5, emerging sequentially as the globally dominant variant. In August 2022, BA.5 was the most common variant circulating worldwide. In India, the proportion of BA.5 infections increased in May 2022, but the proportion of BA.2.75 (a variant of the BA.2 sub-lineage) has increased since June 2022, suggesting that the transmissibility of BA.2.75 may be higher than that of BA.5 (1). Therefore, BA.2.75 has recently been recognized as a variant of concern (VOC) lineage under monitoring. Moreover, in December 2022, BQ.1.1 (a variant of the BA.5 sub-lineage) has been becoming more dominant among Omicron sub-lineages, especially in Europe and the United States (https://cov-spectrum.org). Compared to the SARS-CoV-2 ancestral strain, the BA.1 virus has more than 30 amino acid mutations in the spike protein, including insertions and deletions. The BA.2 spike protein differs from the BA.1 spike protein at 27 amino acid positions, whereas the BA.5 spike protein differs from the BA.2 spike protein by four amino acids, including a L452R mutation. BA.2.75 differs the from BA.2 spike protein at nine amino acid positions, including K147E, W152R, F157L, I210V, and G257S, which are located in the N-terminal domain, and G339H, G446S, N460K, and R493Q, which are located in the receptor-binding domain (RBD). BQ.1.1 differs from BA.5 spike protein at R346T, K444T, and N460K.

Because of the numerous mutations that have accumulated in the spike protein of Omicron variants, Omicron variants are antigenically distinct from the ancestral strain and have the capacity to evade immunity introduced by a primary series of the first-generation COVID-19 vaccine, including COVID-19 mRNA vaccine containing ancestral spike antigen alone. Among Omicron sub-lineages, BQ.1.1 shows the greatest immune evasion against serum neutralization (2-4). In addition, it has been shown that immunity provided by booster vaccination with first-generation vaccines or post-vaccination breakthrough infection can partially protect against Omicron variant infection. However, the surge in SARS-CoV-2 infections has not stopped, even in areas with high booster vaccine uptake, such as Japan. This situation suggests that first-generation COVID-19 vaccines have limited effectiveness at controlling the COVID-19 pandemic, and highlights the need for implementation of the second-generation booster vaccines containing Omicron antigen with improved effectiveness against Omicron variants. Second-generation booster vaccines should induce broad-spectrum protective immunity against all SARS-CoV-2 variants, including Omicron sub-lineages. However, there are many unanswered questions regarding how to induce high-quality immunity that suppresses SARS-CoV-2 variants with distinct antigenicity. A better understanding of the immune response to SARS-CoV-2 variant infection could facilitate the development of better vaccine designs. Specifically, understanding the immune response generated by breakthrough infection or reinfection, which is infection in the presence of pre-existing SARS-CoV-2 immunity due to vaccination or prior infection, respectively, might help to design better booster vaccine antigens (5).

Recently, BA.1 and BA.2 breakthrough infections in individuals vaccinated with COVID-19 mRNA vaccines were found to increase broad serum neutralizing activity against Omicron BA.1, BA.2, and prior VOCs at levels comparable to those of the ancestral strain (6-9). In individuals vaccinated with COVID-19 mRNA vaccines, BA.1 breakthrough infection increases memory B cells primarily for conserved epitopes that are broadly shared among variants and generates robust serum cross-neutralizing activity (9). Notably, convalescent serum samples from individuals with breakthrough infections have higher variable neutralizing activity against Omicron sub-lineages than serum samples of booster vaccination recipients (6-8, 10-15). Furthermore, the exposure interval between vaccination and infection influences the induction of serum cross-neutralizing antibodies against BA.1, with a longer exposure interval contributing to greater induction of serum cross-neutralizing antibodies (12, 16, 17). Unlike booster vaccination, in which the dosing interval between vaccinations is controlled, the exposure interval between vaccination and breakthrough infection is not controlled, resulting in individuals with breakthrough infections having a variable serum neutralizing response to SARS-CoV-2 variants. Therefore, it is difficult to compare the impact of breakthrough infections during different epidemic periods on the serum neutralizing response against the SARS-CoV-2 variants. When the ability to induce serum neutralizing responses through breakthrough infection with Omicron variants and prior VOCs is compared, individuals with breakthrough infections with prior VOCs may have had a shorter exposure interval between vaccination and infection than those with Omicron breakthrough infections, resulting in a lower ability to induce serum cross-neutralizing responses.

In this study, cross-neutralizing activity against SARS-CoV-2 variants, including the Omicron sub-lineages BA.1, BA.2, BA.5, BA.2.75, and BQ.1.1, was assessed using serum samples from individuals with breakthrough infections and booster vaccine recipients before or during the Omicron epidemic. Furthermore, Bayesian modeling was used to correct for the influence of different exposure intervals to enable estimation of the saturated serum cross-neutralizing responses against SARS-CoV-2 variants induced after breakthrough infection with the ideal exposure interval between vaccination and breakthrough infection.

## Results

### Antigenicity of SARS-CoV-2 Omicron sub-lineages in serum samples from individuals with breakthrough infections and booster vaccine recipients

We collected serum samples from individuals with breakthrough SARS-CoV-2 infections during the Omicron BA.1 wave and individuals who had received a booster dose of the first-generation mRNA COVID-19 vaccine containing the ancestral spike antigen alone (SI Appendix, Table S1, and Fig. S1; Material and Methods). Individuals with a history of a COVID-19 diagnosis and positive anti-nucleoprotein (N) antibodies after the second vaccination were defined as having breakthrough infections, whereas individuals who had received three doses of vaccine and did not have a COVID-19 diagnosis or positive anti-N antibodies were defined as booster vaccine recipients in this cohort (Fig. 1A). Similarly, we used serum samples from individuals who had breakthrough infections during the pre-Omicron wave, as reported previously (SI Appendix Fig. S1) (12, 16). Age, exposure interval between the first and second doses of vaccine, and time since the last vaccination were comparable between the three exposure groups (SI Appendix Table S1). Anti-spike (S) antibody titers were highest in individuals with Omicron breakthrough infections, and lowest in individuals with pre-Omicron breakthrough infections (Fig. 1A).

**Fig 1.**
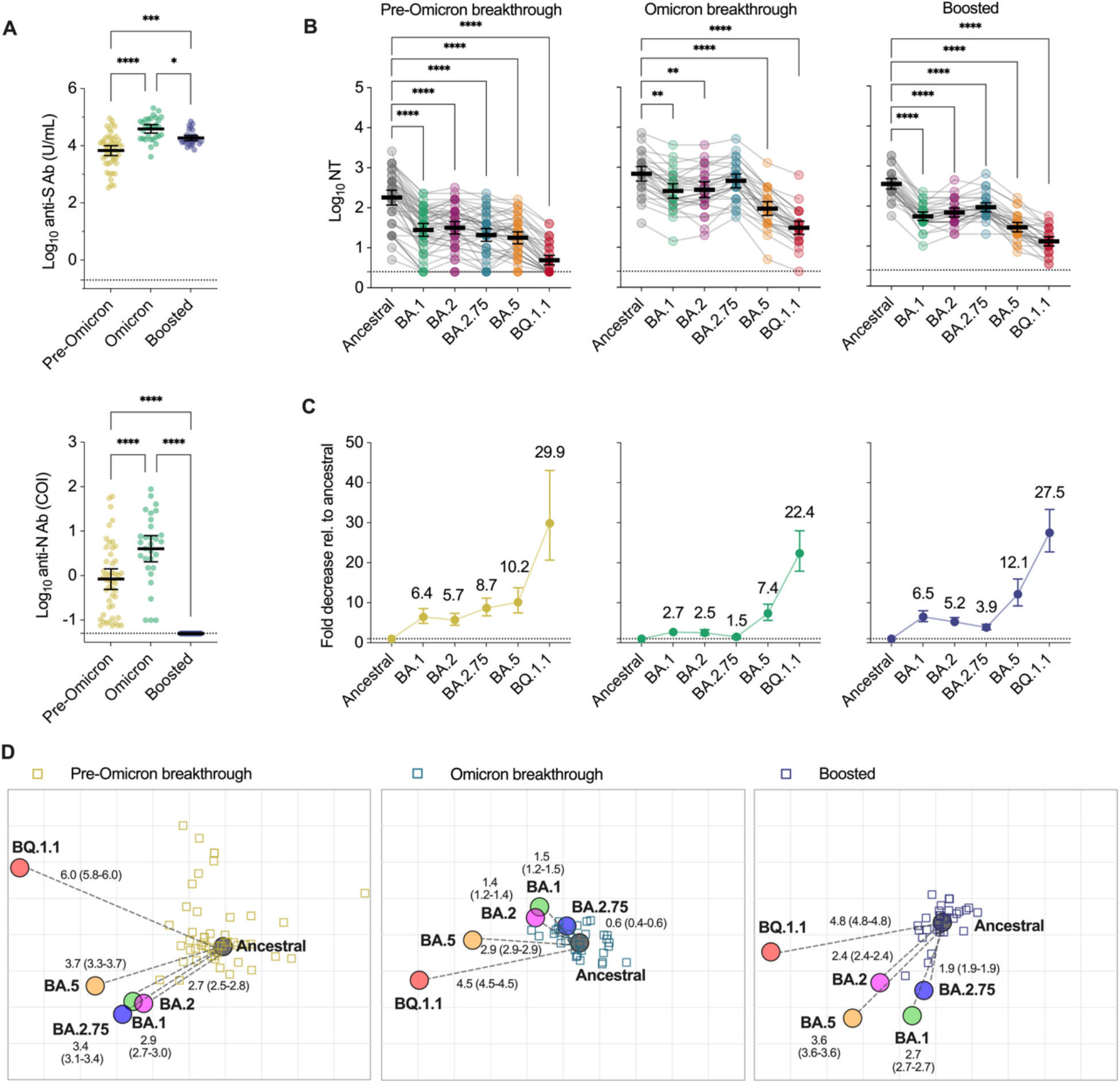
Antigenicity of SARS-CoV-2 Omicron sub-lineages in serum samples from individuals with breakthrough infections and booster vaccine recipients. (A) Anti-spike (S) and anti-nucleoprotein (N) antibody titers in serum samples of individuals with pre-Omicron or Omicron breakthrough infections, and booster vaccine recipients (boosted). The titers were compared using the one-way analysis of variance with the Tukey test. (B) The neutralization titers (NTs) against variants of SARS-CoV-2 live viruses. Data from the same serum sample are connected with lines, and the mean ± 95% confidential interval of each serum titer is presented. The titers between the ancestral and variants were compared using one-way ANOVA with Dunnett’s test. (C) The fold decrease of the NTs against Omicron sub-lineages relative to the NT against ancestral strain. The geometric mean ± 95% confidential interval of each serum sample is shown. (D) Antigenic cartography of each serum source for individuals with pre-Omicron/Omicron breakthrough infections and booster vaccine recipients. The variants are shown as circles and serum samples are indicated as squares. Each square corresponds to a serum sample from one individual. Colors represent the serum source. Each grid square (1 antigenic unit) corresponds to a two-fold dilution in the serum sample used in the neutralization assay. Antigenic distance is interpretable in any direction. The median (95% confidence interval) of the distance from the ancestral strain on the map is shown using gray dotted lines. Statistical significance: \**p*<0.05, \*\**p* <0.01, \*\*\**p*<0.001, \*\*\*\**p*<0.0001.

Neutralization titers (NTs) were determined using live virus-based assays (Fig. 1B). Serum samples from individuals with pre-Omicron breakthrough infections and booster vaccine recipients had uniformly lower NTs against Omicron sub-lineages BA.1, BA.2, BA.2.75, and BA.5, than those against the SARS-CoV-2 ancestral strain (Fig. 1B and 1C). Conversely, serum samples from individuals with Omicron breakthrough infections had high NTs to BA.1, BA.2, and BA.2.75, within 3-fold of the ancestral strain, but 7.4-fold lower NTs to BA.5 (Fig. 1B and 1C). The BQ.1.1 variant had the highest immuno-evasion ability among individuals in the three exposure groups, resulting in a more than 20-fold decrease in the NT relative to that against the ancestral strain (Fig. 1C).

To obtain an overall picture of the antigenicity of Omicron sub-lineages in serum samples from each exposure group, we calculated the positions of antigens and serum samples on antigenic maps based on the difference in NTs (Fig. 1D). Antigenic distances from the ancestral strain to BA.5 and BQ.1.1 evaluated by using all serum samples were further than the distances from the ancestral strain to BA.1, BA.2 and BA.2.75 (Fig. S2). The antigenic distances of BA.1, BA.2, BA.2.75, BA.5, and BQ.1.1 from the ancestral strain in the pre-Omicron breakthrough infection and booster vaccine recipient groups were longer than those in the Omicron breakthrough infection group. In addition, the distance between BA.2.75 and the ancestral strain in the serum samples of the Omicron breakthrough infection and booster vaccine recipient groups was closer than that in the pre-Omicron breakthrough infection group, indicating that BA.2.75 probably exhibited different antigenicity among the three exposure groups. Notably, in serum samples from individuals in the three exposure groups, the antigenic distance between BQ.1.1 and the ancestral strain ranged from 4.5 to 6.0, indicating that BQ.1.1 is the Omicron sub-lineage that is the most antigenically distinct from the ancestral strain. Taken together with Figure 1C, the serum samples of the Omicron breakthrough infection group showed broader breadth of cross-neutralizing potency than those of the pre-Omicron breakthrough infection group.

### Estimating the serum neutralization responses against SARS-CoV-2 Omicron sub-lineages corrected for the influence of different exposure intervals from the second vaccination to the third exposure

A longer time between the second vaccination and third exposure, within a range of approximately 120 days, is necessary to induce broader cross-neutralizing potency in serum samples from individuals with pre-Omicron breakthrough infection, probably because memory B cell affinity maturation occurs during this period (12, 18, 19). The optimal interval between the second vaccination and booster dose has not yet been determined (WHO, https://www.who.int/news/item/17-05-2022-interim-statement-on-the-use-of-additional-booster-doses-of-emergency-use-listed-mrna-vaccines-against-covid-19).

Generally, an interval of 4 to 6 months after the second vaccination could be considered. In Japan, a vaccination interval of at least 3 months is recommended (https://www.mhlw.go.jp/stf/covid-19/booster.html). This vaccination strategy and the periods of pre-Omicron and Omicron waves resulted in distinct exposure intervals among the three exposure groups (Fig. 2A; SI Appendix, Fig. S1, and Table S1). To complement the missing intervals in each exposure history group, we used a Bayesian hierarchical model to estimate the serum neutralizing responses against SARS-CoV-2 variants induced with different antigen exposures and intervals, and estimated the saturated neutralizing responses against SARS-CoV-2 variants with the ideal exposure interval in each exposure history group (Figs. 2 and 3). The overall trend for booster vaccine recipients and individuals with breakthrough infections showed that the interval to saturate the neutralizing response was different for each variant, and that saturating the neutralizing responses against Omicron sub-lineages required a longer exposure interval than those against the ancestral strain (Fig. 2A). To evaluate differences in the exposure interval required to saturate the neutralizing response for each SARS-CoV-2 variant, the probability densities of the estimated number of days to 90% saturated NTs were calculated (Fig. 2B). The medians of the densities of the ancestral strain, and BA.1, BA.2, BA.2.75, BA.5, and BQ.1.1 variants were 34, 74, 71, 106, 95, and 128 days after the third exposure, respectively. This finding suggests that the exposure interval for inducing saturated cross-neutralizing potency against Omicron sub-lineages is longer than that against the ancestral strain, with variants that are more antigenically distant from the ancestral strain requiring a longer period. Additionally, the vaccination-infection intervals in most of the individuals with pre-Omicron breakthrough infections were shorter than the median of the estimated number of days to 90% saturated NTs to Omicron sub-lineages, and these individuals with pre-Omicron breakthrough infections experienced infection without an exposure interval sufficient to acquire cross-neutralizing antibodies. Thus, it is essential to estimate saturated cross-neutralizing potency with the ideal exposure interval to accurately assess the differences in cross-neutralization responses due to varying exposure antigens in breakthrough infections, to avoid bias due to the exposure interval.

**Fig 2.**
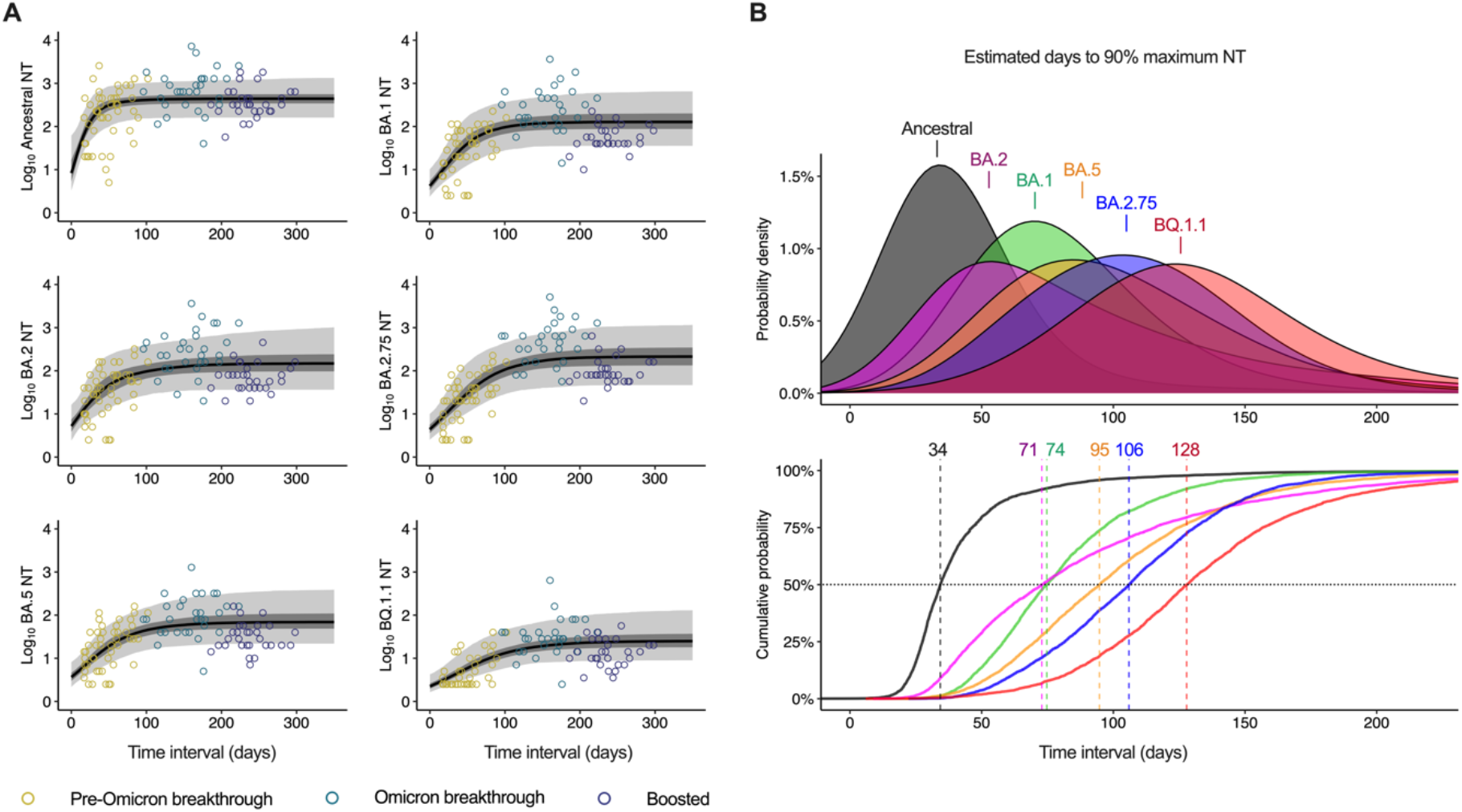
Estimated cross-neutralizing potency against SARS-CoV-2 Omicron sub-lineages of serum samples during the vaccination to third exposure interval in individuals with pre-Omicron/Omicron breakthrough infections or booster vaccination. (A) Estimated increase in the neutralization titers (NTs) in serum samples of individuals with breakthrough infections or booster vaccination (boosted) during the interval from the second vaccination to the third exposure (breakthrough infection or booster vaccination). The measured NTs (circle) and the dynamics estimated by the Bayesian model (posterior median, line; 95% credible interval, light gray ribbon; 50% credible interval, dark gray ribbon) are shown. (B) Estimated exposure interval to 90% saturated NT against SARS-CoV-2 Omicron variants during the period from the second vaccination to the third exposure. The probability density (upper panel, area), the cumulative probability (lower panel, line), and the median time in days (lower panel, dotted line) are shown.

**Fig 3.**
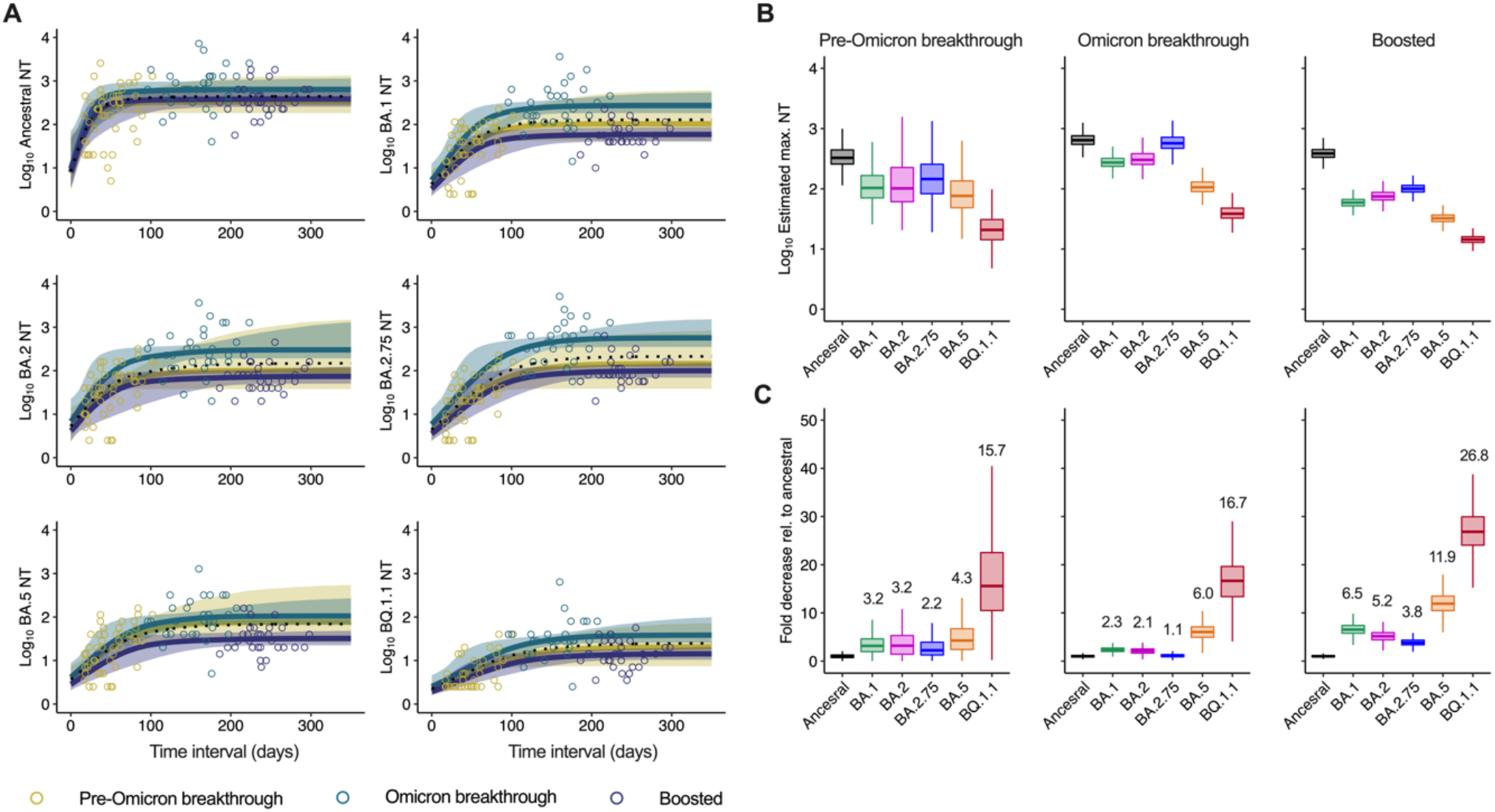
Estimates and comparisons of saturated cross-neutralizing potency of serum samples against SARS-CoV-2 Omicron sub-lineages in individuals with pre-Omicron/Omicron breakthrough infections or booster vaccination. (A) Estimated increases in the neutralization titers (NTs) in serum samples from each participant with pre-Omicron/Omicron breakthrough infection or booster vaccination from the second vaccination to the third exposure (infection or vaccination). The measured NTs (circle) and the dynamics estimated by the Bayesian model (posterior median, line; 95% credible interval, ribbon) are shown. (B) Estimated saturated NTs against SARS-CoV-2 Omicron variants for each exposure group. The posterior median (line), 50% credible interval (box), and 95% credible interval (whisker) are shown. (C) The fold decrease of the NTs relative to the posterior median of NT against the ancestral strain. The median (line), 50% credible interval (box), and 95% credible interval (whisker) are shown. The medians are indicated above the column.

Next, we estimated the saturated neutralizing response to each SARS-CoV-2 variant in each exposure group (Figs. 3A and 3B). In the Omicron breakthrough infection group, the estimated saturated NTs against the ancestral strain, and BA.1, BA.2, BA.2.75, BA.5, and BQ.1.1 variants were clearly higher than those in the booster vaccination group (SI Appendix Figs. S3A, and S3B). Similarly, in the Omicron breakthrough infection group, the saturated NTs against the ancestral strain, and BA.1, BA.2, BA.2.75, and BQ.1.1 variants, were higher than those in the pre-Omicron breakthrough infection group (SI Appendix Figs. S3A, and S3B). In contrast, the saturated NTs against the ancestral strain, and BA.1, BA.2, BA.2.75, and BQ.1.1 variants did not differ significantly between the pre-Omicron breakthrough infection group and the booster vaccination group, but the saturated NT against BA.5 was higher in the pre-Omicron breakthrough infection group than that in the booster vaccination group. Notably, even with the ideal interval, the BQ.1.1 variant exhibited the highest immuno-evasion capabilities in all the exposure groups, with a 15.7–26.8-fold decrease in the saturated NTs relative to those against the ancestral strain (Fig. 3B and 3C). This finding suggests that vaccination and prior infection are less likely to induce protective NTs to BQ.1.1 than to other variants.

Finally, we calculated the fold decrease in the saturated NT relative to the median of the ancestral strain to evaluate the breadth of cross-neutralization potency with the ideal interval (Fig. 3B and 3C). Contrary to the measured NTs (Fig. 1C), no clear differences were observed between individuals with pre-Omicron and Omicron breakthrough infections (Fig. 3C; SI Appendix Figs. S3C and S3D). Notably, compared to the booster vaccination group, the Omicron and pre-Omicron breakthrough infection groups showed relatively mild reduction in the saturated NTs against the Omicron sub-lineages, excluding BQ.1.1. These findings suggest that, given an adequate exposure interval, both Omicron and pre-Omicron breakthrough infections induce a broader breadth of serum cross-neutralizing activity than booster vaccination with the ancestral strain antigen.

## Discussion

In this study, we showed that the ideal exposure interval between vaccination and exposure to achieve saturated neutralizing responses differed by variant in individuals with breakthrough infections, and that more antigenically distant variants from the ancestral strain required a longer exposure interval to reach to a saturated neutralizing response. In addition, we also showed that serum samples from individuals with Omicron breakthrough infections had higher saturated neutralizing responses against the ancestral strain and Omicron sub-lineages than those of individuals with booster vaccination or pre-Omicron breakthrough infection.

An exposure interval of more than 128 days was required to induce broad cross-neutralizing activity against the BQ.1.1 variant, which were antigenically distant from the ancestral strain. As stated above, WHO recommends at least 4 months (approximately 120 days) after the second vaccination before booster vaccination, which is comparable to the exposure interval needed to induce broad cross-neutralizing responses. In contrast, the US Centers for Disease Control and Prevention (CDC) recommends at least 2 months (approximately 60 days) between the second and third doses (https://www.cdc.gov/coronavirus/2019-ncov/vaccines/stay-up-to-date.html). Although a shorter vaccination interval during periods of transient surges in COVID-19 cases may benefit to the level of herd immunity, it may be insufficient to induce high levels of cross-neutralizing antibodies covering antigenically distinct variants. Previous studies have shown that in vaccine recipients, higher NTs were induced with a dose interval of 16 weeks (median, 111 days) than with a dose interval of 4 weeks (median, 29 days) between the first and second doses (20), and the NT and vaccination dose interval were positively correlated within approximately 100 days (21). Notably, when the vaccination dose interval between the second and third dose of vaccine was between 206 and 372 days, there was no difference in the neutralizing responses between the shorter and longer interval (22), suggesting that the effect of vaccination dose interval on inducing neutralizing responses was saturated within this period. These increases and saturation of the serum neutralization response dynamics are consistent with our model. The vaccination dose interval that affects the induction of cross-neutralizing responses can be considered equivalent to the vaccination-infection interval in breakthrough infections in terms of the time taken for the antibody affinity maturation derived from memory B cells in germinal centers after mRNA vaccination (19, 23, 24). Memory B cells that recognize Omicron and other variants proliferate after the second vaccination (18, 22), and the third exposure by vaccination or breakthrough infection induces recall and proliferation of memory B cells which recognize the Omicron spike protein (22, 25). As evaluating the effect of a shorter interval between the second and third vaccine doses would be ethically challenging, our model using individuals with breakthrough infection as a surrogate, provides valuable information about the optimal dose interval between the second and third doses of vaccine. Additionally, our model also suggests that to induce higher levels of cross-neutralizing responses, an additional booster vaccination should be considered in individuals with breakthrough infections with a vaccination-infection interval shorter than four months (120 days).

In serum samples of individuals with Omicron breakthrough infections, the saturated neutralizing responses to Omicron sub-lineages were higher than those in serum samples of individuals with pre-Omicron breakthrough infections and booster vaccine recipients vaccinated at the ideal intervals. Several studies have also found higher cross-neutralizing antibody titers to Omicron sub-lineages in individuals with BA.1 breakthrough infection than in booster vaccine recipients and individuals with Delta breakthrough infection (7-9). Furthermore, BA.1 booster mRNA vaccination of mice and macaques inoculated with the ancestral strain mRNA vaccines also induces higher NTs than a booster dose of the ancestral strain mRNA vaccine (26, 27). Similar findings have also been reported in human studies of Omicron BA.1 and ancestral bivalent vaccine recipients (28), suggesting that booster vaccination with the Omicron antigen induces higher levels of cross-neutralizing antibodies. However, the serum neutralizing breadth in individuals with Omicron BA.1 breakthrough infections was estimated to be similar to those of individuals with pre-Omicron breakthrough infections, regardless of the antigenicity of the infecting variant, and broader than those of booster vaccine recipients. These results suggest that breakthrough infections might contribute to the induction of broader cross-neutralizing responses, referred to as hybrid immunity (5). Compared with three doses of vaccine, breakthrough infection with Delta or Omicron BA.1 variants induces higher levels of memory B cells recognizing ancestral spike RBDs (7, 9, 29), and viral load and duration of viral antigen exposure may contribute to enhanced stimulation of memory B cells. Although the frequency of somatic hypermutations in anti-RBD^+^ memory B cells of individuals with breakthrough infections is comparable to that of booster vaccine recipients (7, 29), antibodies isolated from memory B cells in individuals with breakthrough infections show higher cross-neutralizing activity and affinity (29). Together, these reports and our results suggest that breakthrough infection may contribute to increased cross-neutralizing activity and affinity of memory B cells, regardless of the length of the exposure interval.

### Limitations of study

This study has several limitations. First, the number of samples evaluated was relatively small. Second, the possibility that reduced neutralizing activity at the time of breakthrough infection results in efficient viral replication in the upper respiratory tract may contribute to a better antibody response (16), was not evaluated because of the lack of respiratory specimens. Third, our study does not support the idea that breakthrough infection can act as a substitute for booster vaccination because natural infection can cause long-term complications and is particularly dangerous for vulnerable individuals. Fourth, this study did not include any individuals with a second booster dose of vaccine or breakthrough infection after the first booster dose of vaccine. Finally, our investigation did not evaluate the actual risk of reinfection by SARS-CoV-2 in individuals with a history of breakthrough infection, although there is evidence that NTs are correlated with protection against ancestral strains and different variants (30-32).

## Conclusion

In conclusion, estimating serum cross-neutralizing responses in individuals with breakthrough infection using Bayesian modeling to compensate for the effect of varying exposure intervals, revealed the ideal dose interval and fairly compared the impact of breakthrough infection on breadth of cross-neutralizing responses by variants with distinct antigenicity and epidemic timing. Our results highlight that optimizing the dose interval is critical for maximizing the breadth of cross-neutralizing activity elicited by booster vaccines, with or without Omicron antigens. Understanding how breakthrough infection increases the neutralization breadth would significantly contribute to the development of next-generation COVID-19 booster vaccines covering emerging variants of SARS-CoV-2.

## Materials and Methods

### Participants and sampling

The characteristics of the participants are listed in SI Appendix Table S3 and summarized in SI Appendix Table S1. Serum samples collected 7 to 30 days after the last vaccination were used in the study. The serum samples of booster vaccine recipients and patients with Omicron breakthrough infections were obtained from residual samples of a national seroprevalence survey conducted in Japan from February to March 2022 (peak of the Omicron-dominant period) (SI Appendix Fig. S1) (33). In Japan, the BNT162b2, mRNA-1273, and AZD1222 vaccines have been approved for use since February 2021. Participants received the primary series (doses 1 and 2) at the intervals recommended by the manufacturers. The rollout of the mRNA booster (third) dose was initiated in December 2021, and individuals became eligible 6 to 7 months after the second dose, depending on local availability. Booster vaccinee sera were collected from individuals who had received three doses of BNT162b2 mRNA vaccine, had no history of SARS-CoV-2 infection, and had no detectable anti-nucleoprotein (N) antibody.

Sera from individuals diagnosed with SARS-CoV-2 infection and positive for anti-N antibodies during the Omicron-dominant period following two doses of BNT162b2 or mRNA-1273 mRNA vaccine were used as Omicron-breakthrough infection sera. Based on the date of infection, the majority of the cases of breakthrough infection were probably caused by the Omicron BA.1 and BA.2 lineages, with BA.1, accounting for more than 90% of the cases (SI Appendix Fig. S1). The Omicron-breakthrough infection sera (n= 30) in this study were collected 7 to 30 days after diagnosis of SARS-CoV-2 infection. An equal number of booster vaccine recipients were selected using optimal pair matching based on propensity scores calculated according to age and days since the last exposure (SI Appendix Fig. S1).

Serum samples from patients with pre-Omicron breakthrough infections were obtained as described previously (12, 16). Briefly, pre-Omicron breakthrough infection was defined according to a positive SARS-CoV-2 RNA or antigen test result on a respiratory specimen collected ≥14 days after the second vaccine dose. Demographic information, vaccination status, and respiratory samples for determining the infecting variant were collected as part of the public health activity led by the Japan National Institute of Infectious Diseases (NIID) under the Infectious Diseases Control Law, and the data were published on the NIID website in order to meet statutory reporting requirements. Serum samples obtained from individuals with breakthrough infections were collected concurrently for clinical testing provided by the NIID (with patient consent), and neutralization assays were performed using residual samples as a research activity with ethics approval from the Medical Research Ethics Committee of NIID and informed consent.

To examine neutralization, the serum samples were heat-inactivated at 56°C for 30 min before use. The median dose interval between the first and second vaccine doses for individuals with breakthrough infections and booster vaccine recipients was 21 days (SI Appendix Table S1).

### Ethical statement approval

All samples, protocols, and procedures were approved by the Medical Research Ethics Committee of NIID (approval numbers 1178, 1275, 1312, and 1510).

### SARS-CoV-2 virus

We used the SARS-CoV-2 ancestral strain WK-521 (lineage A, GISAID ID: EPI_ISL_408667), Omicron BA.1 variant TY38-873 (lineage BA.1, GISAID: EPI_ISL_7418017), Omicron BA.2 variant TY40-158 (lineage BA.2.3, EPI_ISL_9595813), Omicron BA.5 variant TY41-702 (lineage BA.5, GISAID: EPI_ISL_ 13241867), Omicron BA.2.75 variant TY41-716 (lineage BA.2.75, GISAID: EPI_ISL_13969765), and Omicron BQ.1.1 variant TY41-796 (lineage BQ.1.1, GISAID: EPI_ISL_15579783) in this study. These variants were isolated using VeroE6/TMPRSS2 cells at NIID with ethics approval provided by the Medical Research Ethics Committee of NIID (#1178). More specifically, viruses belonging to the Omicron lineage were isolated at NIID using VeroE6/TMPRSS2 cells on respiratory specimens collected from individuals screened at airport quarantine stations in Japan and transferred to NIID for whole-genome sequencing.

### Cells

VeroE6/TMPRSS2 cells (JCRB1819, Japanese Collection of Research Bioresources Cell Bank) were maintained in low-glucose Dulbecco’s modified Eagle’s medium (DMEM) containing 10% heat-inactivated fetal bovine serum (FBS), 1 mg/mL geneticin, and 100 U/mL penicillin/streptomycin at 37°C supplied with 5% CO_2_.

### Electrochemiluminescence immunoassay (ECLIA)

Antibody titers for the ancestral spike (S) RBD and nucleoprotein (N) were measured using Elecsys Anti-SARS-CoV-2 S and Elecsys Anti-SARS-CoV-2 kits according to the manufacturers’ instructions.

### Live virus neutralization assay

Live virus neutralization assays were performed as described previously (12, 19). Briefly, serum samples were serially diluted (in two-fold dilutions starting from 1:5) in high-glucose DMEM supplemented with 2% FBS and 100 U/mL penicillin/streptomycin and mixed with 100 median tissue culture infectious dose (TCID_50_) SARS-CoV-2 viruses, followed by incubation at 37°C for 1 hour. The virus-serum mixtures were placed on VeroE6/TMPRSS2 cells seeded in 96-well plates and cultured at 37°C with 5% CO_2_ for 5 days. The cells were then fixed with 20% formalin and stained with crystal violet solution. NTs were defined as the geometric mean of the reciprocal of the highest sample dilution that protected at least 50% of the cells from a cytopathic effect, using two to four multiplicate series. Because of the limited volume of serum samples from individuals with breakthrough infections, this assay was performed only once. All experiments using authentic viruses were performed in a biosafety level 3 laboratory at NIID.

### Antigenic cartography

Antigenic maps based on NTs against SARS-CoV-2 pseudoviruses were created using the *Racmacs* R function with 2,000 optimizations, with the minimum column basis parameter set to “80” (34, 35). Each grid square (1 antigenic unit) corresponded to a two-fold dilution in the neutralization assay. The median the antigenic distances from the ancestral strain and 95% confidence intervals of were calculated according to the Pythagorean theorem using the coordinates of the antigenic maps in the optimization steps.

### Statistical analysis of measured antibody titers

Data analysis and visualization were performed using GraphPad Prism 9.3.1 (San Diego, CA, USA) and R 4.1.2 (https://www.r-project.org/). Measurements below the detection limit were recorded as half the detection limit. One-way analysis of variance with Dunnett’s test or Tukey’s test were used to compare the antibody titers. Statistical significance was set at p < 0.05.

### Estimating saturated cross-neutralizing titers against SARS-CoV-2 variants

For each exposure group, we estimated the NT and time of vaccination using a Bayesian hierarchical model. The log10 NT after breakthrough infection or booster vaccination was described using a three-parameter logistic model for each exposure interval between the second vaccination and the third exposure (vaccination or breakthrough infection). We inferred population means (μ_v_) separately for NTs against the ancestral strain, and BA.1, BA.2, BA.2.75, BA.5, and BQ.1.1 variants. We used a hierarchical structure to describe the distribution of µ_*hv*_ for each exposure group. Arrays in the model index over one or more indices: H=3 exposure history *h*; N=108 participants *n*; V=6 target viruses *v*. The model was as follows:

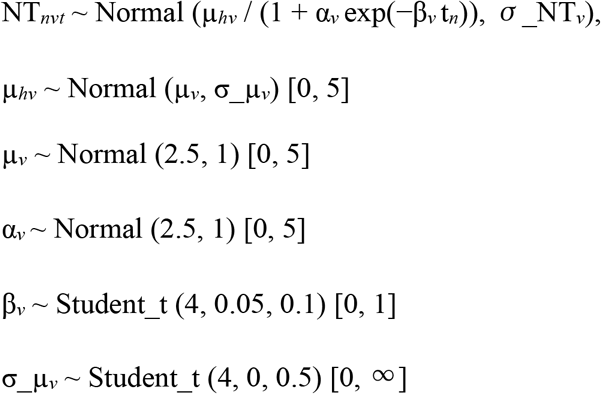

The values in square brackets denote the truncation bounds of the distributions. The explanatory variable was time, *t*_*n*_, and the outcome variable was NT_*nvt*_, which represented the NTs against the target virus *v* in participant *n* at time *t*. A non-informative prior was set for the standard distribution σ_NT_*v*_. The parameters α_*v*_ and β_*v*_ controlled the intercept and the steepness of the logistic function, respectively. The mean parameter for NTs against target virus *v* according to the exposure history *h*, µ_*hv*_, was generated from a normal distribution with hyperparameters of the mean, µ_v_, and standard deviation, σ_µ_*v*_. For the distribution generating β_*v*_ and σ_µ_*v*_, we used a Student’s t distribution with four degrees of freedom, instead of a normal distribution, to reduce the effects of outlier values of β_*v*_ and σ_µ_*v*_.

The exposure interval of days to 90% saturated NTs against each virus (tSNT90_*v*_) was calculated according to the parameters α_*v*_ and β_*v*_ as follows:

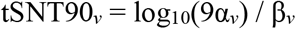

Parameter estimation was performed using a Markov chain Monte Carlo (MCMC) approach implemented in rstan 2.26.1 (https://mc-stan.org). Four independent MCMC chains were run with 5,000 steps in the warm-up and sampling iterations, with subsampling every five iterations. We confirmed that all estimated parameters showed <1.01 R-hat convergence diagnostic values and >500 effective sampling size values, indicating that the MCMC runs were convergent. The fitted model closely replicated the observed NT increases in each exposure group (Figs. 2A and 3A). The above analyses were performed using R 4.1.2 (https://www.r-project.org/). Information on the estimated means of saturated NTs against SARS-CoV-2 variants is summarized in SI Appendix Table S2.

### Data and code availability

Raw data and the code used to estimate the increase in NTs during the interval from the second vaccination to the third exposure are provided in SI Appendix Table S3 and the GitHub repository (https://github.com/ShoMiyamo/VaxInfectionInterval).

## Data Availability

All data produced in the present study are available upon reasonable request to the authors.
The code used to estimate the increase in NTs during the interval from the second vaccination to the third exposure are provided at the GitHub repository.

https://github.com/ShoMiyamo/VaxInfectionInterval

## Acknowledgments

We thank Akiko Sataka, Asato Kojima, Izumi Kobayashi, Yuki Iwamoto, Yuko Sato, Milagros Virhuez Mendoza, Noriko Nakajima, Kenta Takahashi, and Emi Taeda at NIID for their technical support; Jumpei Ito at the University of Tokyo for technical advice on Bayesian modeling; and the healthcare facilities, local health centers, and public health institutes for their contribution in providing us with patient information and samples on pre-Omicron breakthrough cases as listed previously (12). We also thank the Miyagi, Tokyo, Aichi, Osaka, and Fukuoka prefecture governments for their support in implementing the study; staff members at the Survey Research Center and Mitsubishi Research Institute for their administrative and technical assistance; and GISAID for the platform to share and compare our data with data submitted globally. This work was supported by a Japanese Society for the Promotion of Science Grants-in-Aid for Scientific Research (JSPS KAKENHI) grant 21K20768 (to SMi), by Ministry of Health Labour and Welfare (MHLW) grants 20HA2001 (to TS), and 21HA2005 (to TS), and by Japan Agency for Medical Research and Development (AMED) grants JP21fk0108104 (to TS), JP22fk0108637 (to TS), and JP22fk0108141(to TS).

## Supplementary Figures

**Fig S1.**
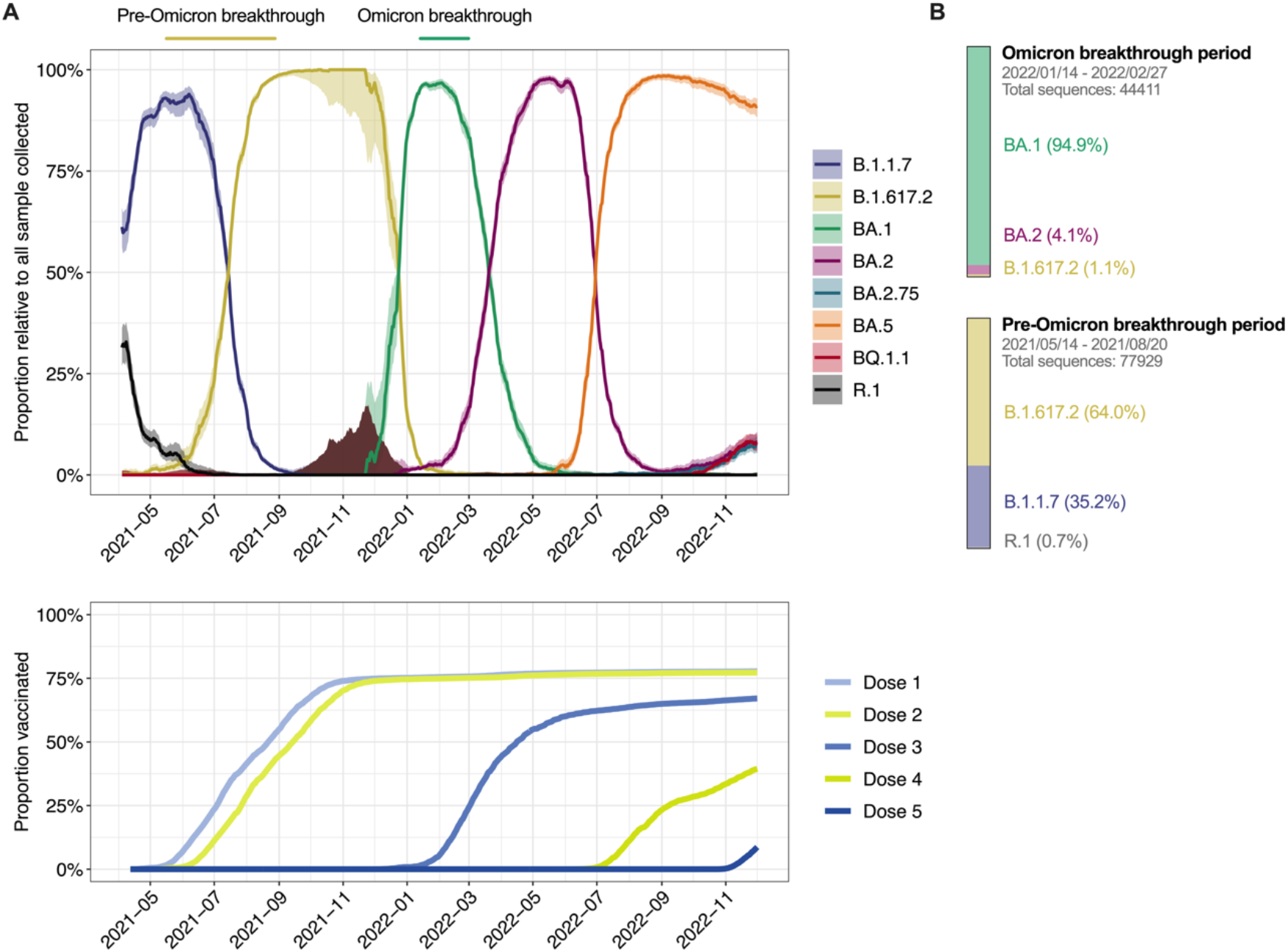
Epigenomic dynamics of SARS-CoV-2 lineages in Japan. (A) The proportion of each variant relative to all sample sequences collected (line) with the 95% confidential interval (ribbon) (upper left), and the vaccination coverage of the population in Japan (lower left) are shown. Among individuals with pre-Omicron and Omicron breakthrough infections, the dates of infection are shown at the top. (B) The proportion of SARS-CoV-2 lineages according to the date of each breakthrough infection. Based on the GISAID database (https://platform.epicov.org), CoV-Spectrum (https://cov-spectrum.org) provided the percentage, the 95% confidence interval, and the number of sequences. Digital Agency, Japan (https://info.vrs.digital.go.jp/dashboard) provided the data on the proportion vaccinated.

**Fig S2.**
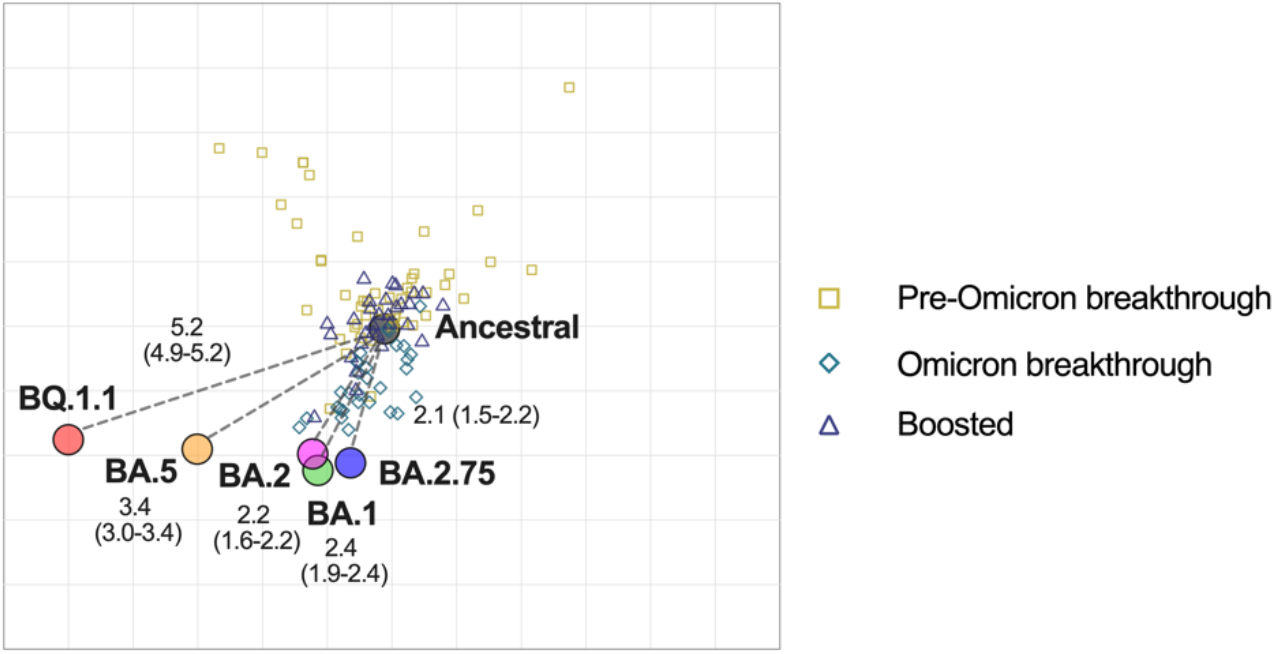
Antigenicity of SARS-CoV-2 Omicron sub-lineages in heterogeneous serum samples from individuals with breakthrough infections and booster vaccine recipients. Antigenic cartography of serum sources for individuals with pre-Omicron/Omicron breakthrough infections and booster vaccine recipients. The variants are shown as circles and serum samples are indicated as squares, diamonds, and triangles. Each square, diamond, and triangle corresponds to a serum sample from one individual. Each grid square (1 antigenic unit) corresponds to a two-fold dilution in the serum sample used in the neutralization assay. Antigenic distance is interpretable in any direction. The median (95% confidence interval) of the distance from the ancestral strain on the map is shown using gray dotted lines.

**Fig S3.**
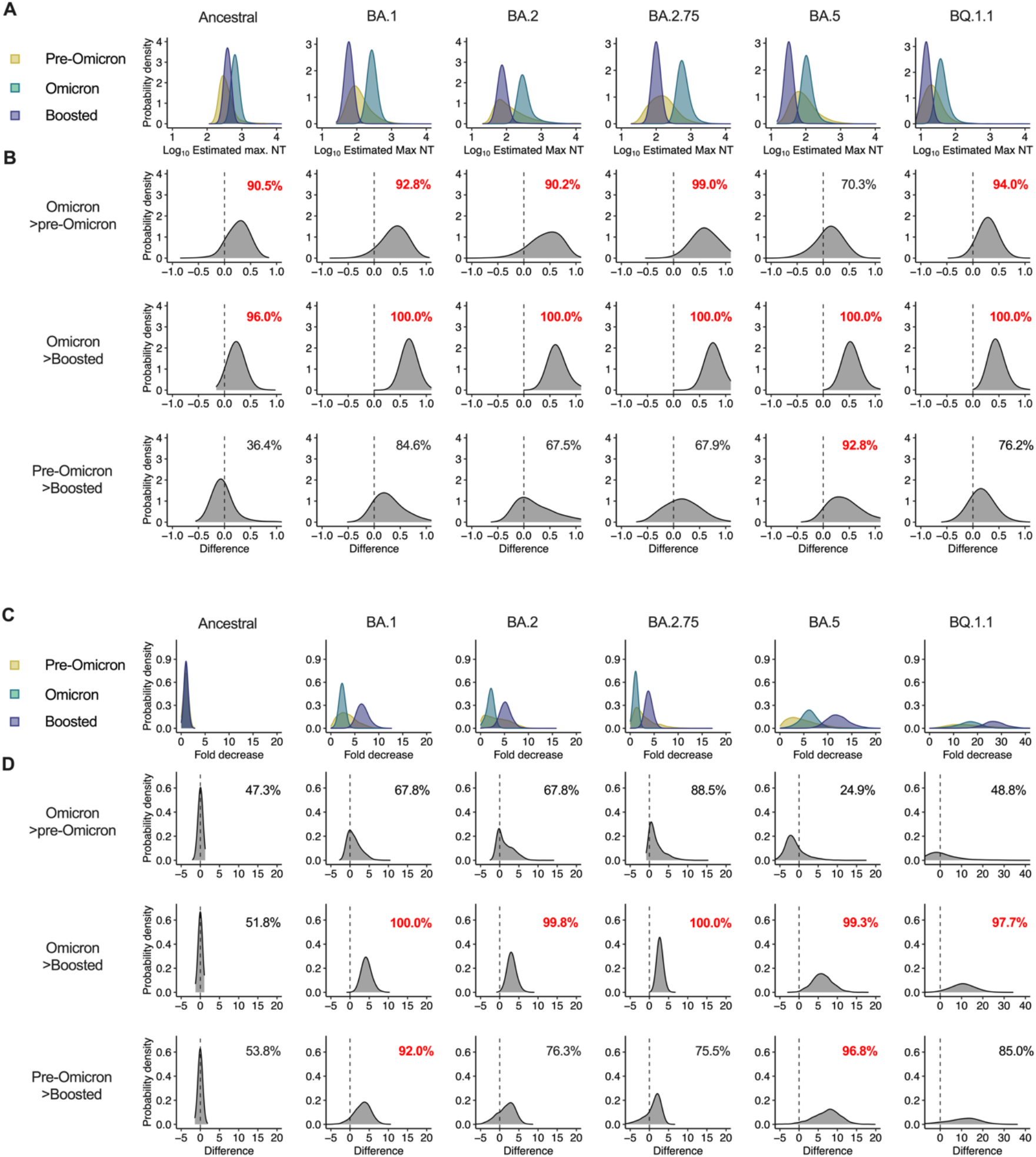
Estimates of the probability that the neutralizing titer and the fold decrease differs according to the exposure history. (A, B) Probability densities of estimated maximum neutralization titers (NTs) against SARS-CoV-2 ancestral strain and Omicron sublineages for each exposure history. (B) Probability density of NT differences between two exposure histories. The indicated probabilities are displayed and highlighted in red if the probability is >90.0%. (C, D) Probability densities of a fold decrease in the NTs relative to the posterior median of NT against the ancestral strain. (D) Probability density of fold decrease differences between two exposure histories. The calculated probabilities are displayed and highlighted in red if the probability is >90.0%.

## Supplementary Tables

**Table S1.** Characteristics of study participants.

|  | Pre-Omicron<br>breakthrough | Omicron<br>breakthrough | Booster<br>vaccination |
| --- | --- | --- | --- |
| **Sample** | Serum | Serum | Serum |
| **N** | 48 | 30 | 30 |
| **Age (y)** | 36 (26, 50) | 48 (38, 54) | 44 (37, 52) |
| **Male sex** | 14 (29%) | 8 (27%) | 4 (13%) |
| **Vaccine (2 doses)** |  |  |  |
| BNT162b2 | 47 (98%) | 19 (63%) | 30 (100%) |
| mRNA-1273 | 0 (0%) | 11 (37%) | 0 (0%) |
| Not listed | 1 (2.1%) | 0 (0%) | 0 (0%) |
| **Dose 1 to dose 2**<br>**interval (days)** | 21 (21, 21) | 21 (21, 28) | 21 (21, 21) |
| **Dose 2 to infection**<br>**(days)** | 44 (30, 62) | 166 (134, 177) | NA |
| **Dose 2 to dose 3**<br>**interval (days)** | NA | NA | 237 (223, 254) |
| **Last exposure to**<br>**collection (days)** | 14 (11, 15) | 20 (14, 24) | 19 (15, 25) |
| **Period of infection** | May. 14 -<br>Aug. 20, 2021 | Jan. 15 -<br>Feb. 27, 2022 | NA |

Median (interquartile range); n (%)

**Table S2.** Means of the maximum neutralization titers against SARS-CoV-2 variants.

| Target virus | Exposure-history | Posterior mean | Posterior 2.5% | Posterior 97.5% | R-hat | Effective sample size |
| --- | --- | --- | --- | --- | --- | --- |
| Ancestral | Total | 2.65 | 2.21 | 3.14 | 1.00 | 2092 |
| Ancestral | Pre-Omicron breakthrough | 2.56 | 2.26 | 3.14 | 1.00 | 1165 |
| Ancestral | Omicron breakthrough | 2.82 | 2.61 | 3.06 | 1.00 | 1039 |
| Ancestral | Booster vaccination | 2.59 | 2.41 | 2.77 | 1.00 | 1736 |
| BA.1 | Total | 2.13 | 1.55 | 2.82 | 1.00 | 3267 |
| BA.1 | Pre-Omicron breakthrough | 2.06 | 1.62 | 2.79 | 1.00 | 3320 |
| BA.1 | Omicron breakthrough | 2.45 | 2.26 | 2.74 | 1.00 | 2468 |
| BA.1 | Booster vaccination | 1.77 | 1.61 | 1.94 | 1.00 | 3368 |
| BA.2 | Total | 2.21 | 1.56 | 3.12 | 1.00 | 1954 |
| BA.2 | Pre-Omicron breakthrough | 2.13 | 1.58 | 3.30 | 1.00 | 1465 |
| BA.2 | Omicron breakthrough | 2.54 | 2.28 | 3.25 | 1.00 | 1291 |
| BA.2 | Booster vaccination | 1.89 | 1.70 | 2.15 | 1.00 | 1882 |
| BA.2.75 | Total | 2.34 | 1.67 | 3.07 | 1.00 | 3758 |
| BA.2.75 | Pre-Omicron breakthrough | 2.18 | 1.59 | 2.95 | 1.00 | 3413 |
| BA.2.75 | Omicron breakthrough | 2.78 | 2.55 | 3.21 | 1.00 | 3165 |
| BA.2.75 | Booster vaccination | 2.00 | 1.84 | 2.18 | 1.00 | 3527 |
| BA.5 | Total | 1.89 | 1.34 | 2.65 | 1.00 | 2978 |
| BA.5 | Pre-Omicron breakthrough | 1.94 | 1.41 | 2.79 | 1.00 | 3036 |
| BA.5 | Omicron breakthrough | 2.05 | 1.83 | 2.46 | 1.00 | 2475 |
| BA.5 | Booster vaccination | 1.51 | 1.35 | 1.70 | 1.00 | 3282 |
| BQ.1.1 | Total | 1.44 | 0.95 | 2.17 | 1.00 | 3140 |
| BQ.1.1 | Pre-Omicron breakthrough | 1.33 | 0.87 | 1.92 | 1.00 | 2655 |
| BQ.1.1 | Omicron breakthrough | 1.63 | 1.41 | 2.08 | 1.00 | 1207 |
| BQ.1.1 | Booster vaccination | 1.16 | 1.02 | 1.35 | 1.00 | 1204 |

## Notes

### Competing Interest Statement

The authors have declared no competing interest.

### Author Declarations

Ethics committee of the National Institute of Infectious Diseases gave ethical approval for this work.

